# APOE4 Allele Frequencies Show Dramatic Variation Across Indian Populations

**DOI:** 10.64898/2026.04.09.26350483

**Authors:** Shweta Ramdas, Bratati Kahali

## Abstract

The *APOE* ε4 allele is the strongest genetic risk factor for Alzheimer’s Disease. However, its distribution across Indian populations is poorly characterized. We analyze *APOE* allele frequencies in 9,524 individuals from 83 distinct populations in the GenomeIndia dataset. ε4 frequencies show large variation across populations within India, ranging from 2.7% to 36.1%, with a median of 11%. Tribal populations have higher ε4 frequencies compared to non-tribal groups, while Tibeto-Burman populations have significantly lower frequencies. One tribal population from the northern coastal highlands has ε4 frequency of 0.36, with 59% of individuals being carriers. ε4 carrier status correlates significantly with lipid phenotypes including LDL, HDL, total cholesterol, and triglycerides. Collectively, these findings reveal exceptional genetic diversity in Alzheimer’s Disease risk across India and have important implications for population-specific screening strategies, genetic counseling, and precision medicine approaches to dementia prevention.

## Introduction

The *APOE* locus is the strongest genetic risk factor for Alzheimer’s Disease (AD), with the ε4 isoform being the strongest risk allele, and the ε2 allele being protective. The e2 allele, with the lowest allele frequency among the three most common isoforms, is protective against AD. The effect size of the APOE4 allele is population-biased, with Europeans showing the largest effect sizes, and African populations showing lower effect sizes.

A few studies (1,2) quantified the impact of ε4 frequencies on cognitive function in India; one in a rural cohort from South India, another (2) in an aging cohort in individuals 45 and older. In this study, we characterize the diversity in genetic variation at the *APOE* locus across 83 populations in India, catalogued in the GenomeIndia project (GI) (3). India has more than 4,600 ethnic groups, and the documented genetic diversity due to endogamy and founder effects can lead to starkly different risk prevalences in each group. Population-stratified analyses can identify communities that are either at risk or protected from the genetic risk of AD.

## Results

### Large variation in ε4 allele frequencies across the country

The median allele frequency of *APOE* ε4 across the GI dataset is 0.11, in line with previous studies (2,4). The alleles follow Hardy-Weinberg equilibrium across the 9,524 analysed samples; 143 individuals are homozygous for the ε4 allele.

However, allele frequencies vary largely across populations, from 0.027 to 0.361 (Figure 1A; Table S1). Consistent with the wide range, *APOE* shows moderate population differentiation within India (multiallelic Weir–Cockerham *F*_*ST*_ = 0.026). ε2 and ε4 allele frequencies across populations are positively correlated (Spearman correlation of 0.27, P = 0.03).

**Figure 1.**
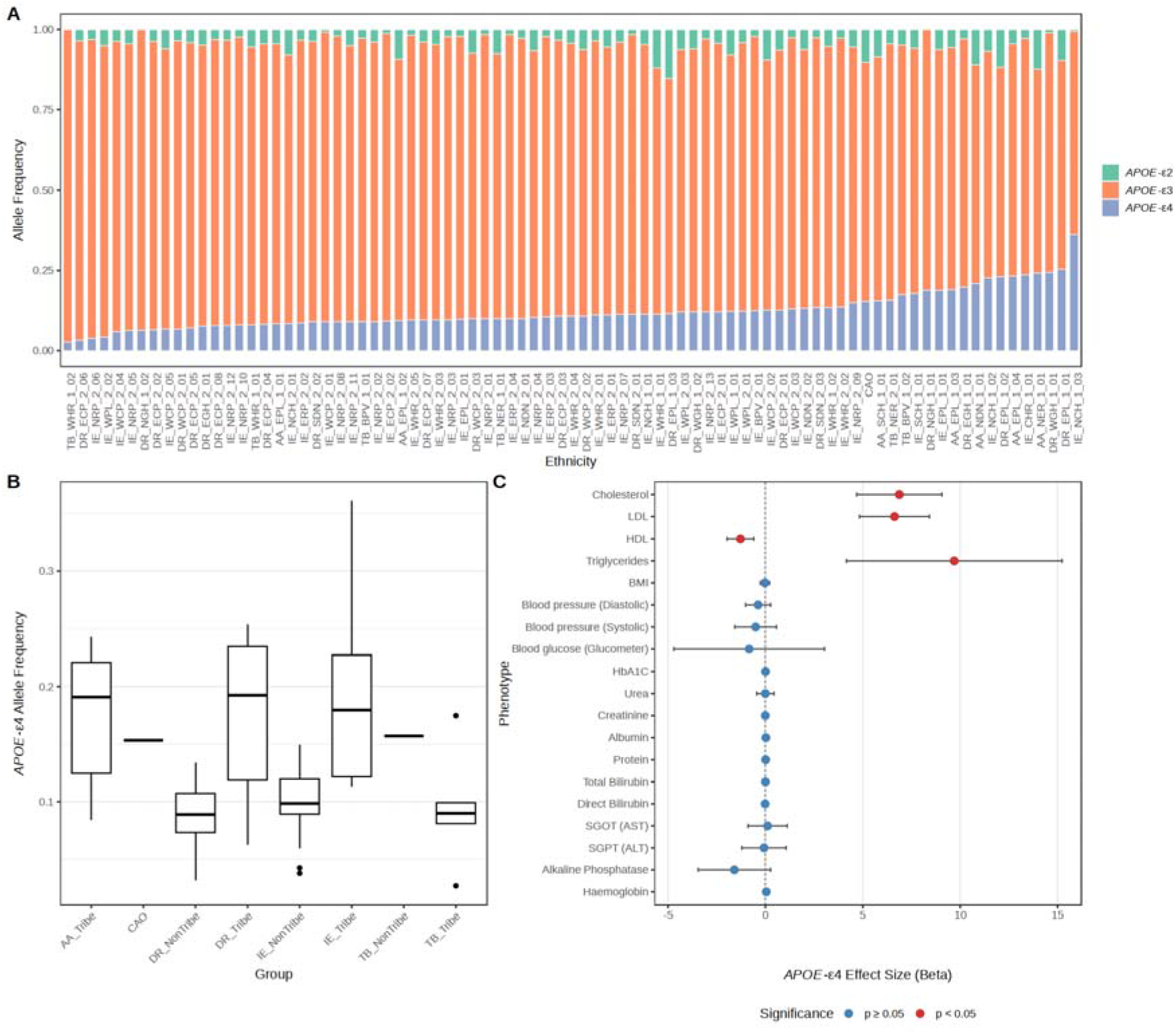
Variation in *APOE* haplotypes across ethnicities in GenomeIndia. **A.** Allele frequencies of ε4 across ethnicities, sorted by increasing ε4 frequency. B. ε4 distributions along different ancestries, and between tribal and non-tribal populations C. Correlations of ε4 carrier status with blood biochemistry traits

A tribal population from the Indo-European linguistic family from the northern coastal highlands (IE_NCH_1_03) has the highest ε4 frequency of 0.36 (confidence interval 0.29-0.44). This population has an ε2 allele frequency of 0.007 (in the bottom 10%), indicating a large genetic load of risk for AD. 59% of sampled individuals in this population are ε4 carriers. Permutation testing shows that this high frequency is unlikely by chance (1/1000 permutations of e4 alleles in 83 populations.) The variant is in HWE in this population.

On the other end of the distribution, population TB_WHR_1_02 from Himachal Pradesh, a Tibeto-Burman tribal population from the Western Himalayan Region (56 sampled individuals) has ε4 allele frequency of 0.027, indicating possibly lower AD genetic burden. Indeed, populations from the Western Himalayan range of Tibeto-Burman ancestry have been reported to have lower AD prevalences (5). The highest frequency of the ε2 allele (0.15) is in a Dravidian tribal population from Odisha (ε4 AF 0.11).

### Association with ancestry and blood biochemistry

Tribal populations have significantly higher ε4 frequencies (beta 0.06, p = 2.2e-07). Tibeto-Burman populations have significantly lower ε4 frequencies (Figure 1B), with no significant differences between Indo-European, Dravidian and Austro-Asiatic groups. After correcting for tribal status and ancestry, biogeography, age, and gender are not correlated with ε4 allele frequencies.

After correcting for tribe status and gender, ε4 carrier status correlates with lipid phenotypes (LDL, HDL, total cholesterol, and triglycerides), with ε4 carriers having higher cholesterol and triglyceride levels (P-value < 0.0026, the Bonferroni threshold) (Figure 1C).

## Discussion

This study reveals unprecedented variation in *APOE* allele frequencies across Indian populations, with implications for both population genetics and clinical practice. The 14-fold difference in ε4 frequencies (2.7%-36.1%) represents one of the largest documented ranges within a single country, highlighting India’s exceptional genetic diversity. To our knowledge, this represents the largest population-resolved survey of *APOE* allele frequencies within India.

The finding that Tibeto-Burman populations have significantly lower ε4 frequencies aligns with epidemiological reports of reduced dementia prevalence in these communities. Conversely, populations with high ε4 frequencies and absent protective ε2 alleles may require enhanced screening and preventive strategies. The strong association between tribal status and elevated ε4 frequencies raises questions about access to neurological care in these communities, which are often geographically isolated.

The correlation between *APOE* ε4 and lipid metabolism observed here is consistent with APOE’s role in cholesterol transport. The population-specific variations in these associations warrant further investigation.

This study has several limitations; the lack of clinical outcome data makes any risk estimates putative. Longitudinal studies with cognitive assessments are needed to confirm the clinical significance of these genetic variations. Additionally, environmental and lifestyle factors that may modulate *APOE* effects were not assessed.

## Methods

### Samples

The underlying genetic data were drawn from the GI dataset (3). Allele frequencies were estimated in 9,524 unrelated samples. Each population was annotated with its linguistic group (Indo-European, Dravidian, Tibeto-Burman, Austro-Asiatic, CAO), tribal status, and geographic region. 9,098 individuals had phenotypic data available, including blood biochemistry, socio-demographic, and anthropometric variables. Variants passed rigorous QC.

### Statistical Analysis

Population-specific allele frequency confidence intervals were calculated using Wilson score intervals for binomial proportions. Population differences were assessed using linear regression models correcting for tribal status, ancestry, and geographic region. Biochemical correlations were analyzed using linear regression with Bonferroni correction (P < 0.0026). Population differentiation at the *APOE* locus was quantified using multiallelic Weir–Cockerham *F*_*ST*_, estimated from individual-level genotypes.

## Data availability

Genotype data are available at the Indian Biological Data Centre.

## Author contributions

S.R. and B.K. designed the study. S.R. analysed data and wrote the manuscript.

## Acknowledgements

We thank all participants who consented to providing their samples for this project.

